# Investigating the association between maternal infection and inflammation and child autistic traits in a large population based cohort study

**DOI:** 10.1101/2025.09.11.25335326

**Authors:** Frederieke A. J. Gigase, Milan Zarchev, Ryan L. Muetzel, Charlotte A. M. Cecil, Luz H. Ospina, Manon H. J. Hillegers, Rebecca Birnbaum, Lot D. de Witte, Veerle Bergink

**Affiliations:** Department of Child and Adolescent Psychiatry/Psychology, Erasmus MC, Rotterdam, the Netherlands; The Generation R Study Group, Erasmus MC, Rotterdam, The Netherlands; Department of Radiology and Nuclear Medicine, Erasmus MC, Rotterdam, the Netherlands; Department of Psychiatry, Icahn School of Medicine at Mount Sinai, New York City, NY, United States; Department of Genetics and Genomic Sciences, Icahn School of Medicine at Mount Sinai, New York City, NY, United States; Department of Human Genetics, Radboud UMC, Nijmegen, the Netherlands; Department of Psychiatry, Radboud UMC, Nijmegen, the Netherlands; Department of Psychiatry, Erasmus MC, Rotterdam, the Netherlands; Department of Obstetrics, Gynecology and Reproductive Science, Icahn School of Medicine at Mount Sinai, New York City, NY, United States

## Abstract

**Objective:** Maternal immune activation during pregnancy has been proposed as a mechanism linking prenatal inflammatory exposures to autism pathogenesis. While preclinical and epidemiological studies suggest a role for maternal inflammation and infection, findings in population-based cohorts are inconsistent. This study examined the associations between multiple prenatal inflammatory exposures and autistic traits, accounting for gene-environment interactions in the general pediatric population.

**Methods:** We leveraged data from 5,075 mother-child dyads participating in Generation R, a population-based pregnancy cohort in the Netherlands. Prenatal inflammatory exposures included 1) maternal serum cytokines; 2) high-sensitivity CRP; 3) self-reported fever during pregnancy; 4) a maternal polygenic score for CRP; and 5) a methylation profile score of CRP in cord blood. Child autistic traits were measured with the Social Responsiveness Scale at mean ages 6 and 13 years. Linear mixed models were applied to estimate associations adjusted for maternal, child and technical covariates. Interaction terms tested whether child polygenic score for autism moderated associations.

**Results:** No significant associations were observed between prenatal inflammatory exposures and autistic traits, both as a continuous measure and above a clinical threshold. No evidence was found for interactions between prenatal inflammatory exposures and the child polygenic score for autism in influencing autistic traits.

**Conclusion:** Our findings suggest that typical fluctuations in maternal inflammation are unlikely to represent a major pathway linking prenatal environment to autism risk. We found no evidence that gene-environment interactions conferred additional risk for autistic traits.

## Introduction

Autism spectrum disorder (ASD) is a neurodevelopmental disorder characterized by repetitive behaviors and deficits in social-emotional interactions (1). Though a categorical diagnosis, dimensionally it represents the extreme end of a continuous distribution of autistic traits observed in the general population. Prevalence of childhood onset ASD among US children has increased recently (2). Studies consistently demonstrate a high heritability of ∼80%, while non-genetic factors may also significantly influence autism risk (1,3,4). Among these, prenatal infections and inflammation, together referred to as maternal immune activation (MIA) – have emerged as biologically plausible environmental contributors to autism pathogenesis.

Epidemiological studies indicate that prenatal infections during pregnancy are associated with a 30– 50% increased risk of autism diagnosis in the child, with stronger associations observed for severe infections and exposures during early gestation (5–8). Preclinical models have provided insight in potential mechanisms, demonstrating that maternal exposure to infections or systemic inflammation can disrupt fetal brain development via IL-6 and IL-17A mediated inflammatory signaling, impairing neurogenesis, cortical lamination, and microglial maturation, and consequentially offspring behavior (9–11). Yet, robust evidence in general population cohorts is lacking. While some studies have linked elevated maternal cytokines (including interleukin (IL)-1, IL-6 and interferon (IFN)-y) to child ASD diagnosis (12,13), others have found no association between prenatal infections or inflammation and child autistic traits (14–16). Given that most children exposed to maternal immune activation do not develop ASD, gene–environment interactions likely modulate susceptibility to immune-mediated neurodevelopmental disruption, with individuals at higher genetic risk being more susceptible to autistic traits following prenatal immune disruptions. Yet, child genetic liability for ASD is rarely examined in MIA research. Moreover, the inconsistent findings among epidemiological, preclinical and population-based studies may in part stem from the heterogeneity of the MIA field, encompassing inflammatory exposures that differ in type, timing, and severity. Differentiating between acute phase reactants and more stable inflammatory proxies will be crucial to clarify the contribution of MIA to ASD risk.

The current study aimed to address these issues by leveraging the Generation R cohort, a large prospective pregnancy study in Rotterdam, the Netherlands. We examined multiple inflammatory exposures across gestation and at birth in relation to autistic traits. While prior research has typically relied on a single exposure, we have leveraged the rich Generation R data to thoroughly compare a broad set of MIA indicator variables in this large general population cohort. These included: 1) maternal serum cytokines (IL-1β, IL-6, IL-17a, IL-23, IFN-γ) and 2) c-reactive protein (CRP), as these samples are taken at standard time points during pregnancy, the measurements reflect the chronic peripheral inflammatory state, also referred to as ‘low-grade inflammation’ (17); 3) self-reported fever, reflecting an acute inflammatory response; 4) maternal polygenic score (PGS) of CRP, reflecting genetic predisposition to chronically elevated CRP; and 5) a methylation profile score (MPS) of cord blood CRP, reflecting stable epigenetic signatures of systemic inflammation at birth. By integrating detailed inflammatory exposures with genetic susceptibility to ASD in a general population sample, this study aims to clarify the role of maternal immune activation in the development of autistic traits and to identify gene-environment mechanisms underlying individual differences in vulnerability.

## Methods

### Study population

This project was conducted within the Generation R Study – a large prospective population-based pregnancy cohort investigating health and development from fetal life onwards. Data collection for the Generation R Study took place between April 2002 and January 2006 at the Erasmus Medical Center, Rotterdam, The Netherlands (18). All participants provided written informed consent. The Medical Ethics Committee of Erasmus MC approved all study procedures. The current study includes all Generation R mother-child dyads with at least one available measurement of blood serum cytokines, fever questionnaire data, and at least one available assessment of child autistic traits. Of the 9,901 mother-child dyads participating in the baseline measurement, 1,678 mothers had no complete data on cytokine and CRP levels at any timepoint and 369 further mothers had no data on self-reported fever. Moreover, 2,723 children did not have SRS data at any timepoint. Finally, we excluded one sibling for each pair based on data completeness (n=57). The final sample included 5,075 mother-child dyads, of which 2,151 were included in analyses on the MPS-CRP.

### Inflammatory exposures

#### HS-CRP & Cytokine Index

Maternal serum samples were collected at two timepoints at median 13.2 weeks gestation (95% range: 9.6-17.1 weeks) and median 20.4 weeks gestation (95% range: 18.5-22.7 weeks) through antecubital venous puncture. Blood samples were processed within three hours of collection, centrifuged for 10 minutes and immediately stored at -80°C. HS-CRP was measured using an immunoturbidimetric assay on the Architect System available through the Department of Clinical Chemistry of the Erasmus MC (Abbot Diagnostics B.V., Hoofddorp, the Netherlands). Cytokines IL-1β, IL-6, IL-17a, IL-23, and IFN-γ were selected based on a pilot study and measured using the Human High Sensitivity T-Helper Cells Custom 5-plex assay from Millipore (Millipore, St. Charles, MO, USA) on the Luminex™ 100 system (Luminex, Austin, TX, USA) by Eve Technologies Corp. (Calgary, Alberta) in 2023, as described previously (17). Cytokines were summarized into a cytokine index, a summary score equal to the first principal component (17). The average of the two timepoints was included as exposure in the main analyses.

#### Fever

Data on prenatal infections was collected during each trimester of pregnancy via a self-report questionnaire. Participants were asked to report whether they experienced a period of fever (>38°C/100.4°F) within the past 2 (second trimester) or 3 months (first and third trimester). A count variable was created, indicating the number of fevers reported throughout the pregnancy (range 0-3).

#### PGS-CRP

The maternal polygenic score (PGS) of CRP reflects a genetic predisposition to chronically elevated CRP levels, thereby capturing a stable liability to low-grade inflammation that is not influenced by transient environmental factors. The PGS-CRP was constructed as described previously (17). In short, maternal samples were used for DNA extraction, as detailed elsewhere (19). Mothers in Generation R were genotyped in two batches using the Illumina Global Screening Multi-Disease Array (GSA-MD) v2 (1,530 mothers in 2019/2020) and GSA-MD v3 (10,491 mothers in 2022) platforms. Both batches underwent extensive quality control checks. A recent large-scale gene-wide association study (GWAS) with 575,531 participants was used to construct a PGS of CRP based on imputed genotypic data (20). The summary statistics were obtained from the GWAS study catalog (ID: GCST90029070). LDpred2 was used to calculate the PGS (21,22). We used a genome-wide BED file with the maternal genotypes, HapMap3 variants with individual LD matrix in blocks and LDpred2-auto (one variant of LDpred2) to automatically estimate p (the proportion of causal variants) and h^2^ (the SNP heritability) from the summary statistics. The calculated score was standardized to z-scores (mean 0, 1 SD) and residualized on the first 10 principal components.

#### MPS-CRP

A methylation profile score of CRP (MPS-CRP) in cord blood serves as an epigenetic proxy of neonatal inflammation, providing a stable measure of systemic immune activation at birth (23). The construction and validation of the MPS-CRP in cord blood was described previously (23). In short, umbilical cord blood was drawn and DNAm profiles were generated either with the Illumina Infinium HumanMethylation450 BeadChip array (Illumina Inc., San Diego, CA) (GenerationR_450k_: n = 1,202) or with the Illumina MethylationEPIC 850 K array (Illumina Inc., San Diego, CA) (GenerationR_EPIC_: n = 949) for a total of 2,151 participants. Normalized, untransformed beta values were used that ranged from 0 (i.e. fully unmethylated) to 1 (i.e. fully methylated) to indicate methylation levels. Seven DNA methylation probes (CpGs) were selected based on their reported association with CRP as well as inflammation-related conditions in a previously published EWAS, performed in adults and based on whole blood (24). Next, the methylation beta value at each probe was multiplied by the corresponding regression betas from the EWAS. The weighted methylation values were then summed into a single MPS-CRP and standardized to z-scores to enable comparison. Analyses were conducted separately for each DNAm array subcohort (GenerationR_450k_ vs GenerationR_EPIC_), after which the results were meta-analyzed using the metafor package (25).

### PGS for ASD

A polygenic score (PGS) for ASD, reflecting the child’s genetic susceptibility for autism, was computed using a clumping and thresholding approach in PRSice2 as described previously (26). In short, genetic single nucleotide polymorphism (SNP) data identified from a GWAS of 18,381 cases and 27,969 controls was used to generate the PGS for ASD (27). The PGS was standardized to z-scores and residualized on the first 10 principal components.

### Assessment of autistic traits

Child autistic traits were measured at mean age 6 and 13 years, using the abbreviated parent-reported Social Responsiveness Scale (SRS; 18-items) (28). Participating parents were asked to respond to an 18-item version of the questionnaire for children on a 4-point Likert scale. The SRS is a validated questionnaire assessing the severity of autism-related traits in the general population, including interpersonal behavior, communication, and stereotypic behavior (28). The abbreviated SRS questionnaire was studied in prior Generation R publications (29), and is highly correlated (r=0.93) to the full SRS with a Cronbach’s alpha value of 0.92. Higher mean scores are indicative of higher levels of autistic traits. Z-scores of SRS mean scores were computed. As part of secondary analyses, scores from the 18-item SRS were gender-weighted to align with recommended cutoffs for population-based screening (1.078 for boys and 1.000 for girls), derived from the suggested thresholds for the full 65-item SRS. Based on these weighted scores, the SRS was dichotomized into clinical versus subclinical autistic traits, with children classified as clinical if their score exceeded the cutoff at any timepoint (28).

### Statistical analysis

#### Main analyses

Mixed-effects linear regression analyses were performed to analyze the association between inflammatory exposures and child autistic traits. Separate regression models were performed for each exposure: 1) A composite cytokine index based on maternal serum IL-1β, IL-6, IL-17a, IL-23, and IFN-γ; 2) Maternal serum CRP; 3) Self-reported fever during pregnancy; 4) Maternal polygenic score (PGS) of CRP; and 5) Methylation profile score (MPS) of cord blood CRP. First, we investigated the average association between each exposure and autistic traits. Models included a random intercept for participant ID to account for repeated observations per person. In addition, timepoint-specific associations were estimated separately for child age at assessment (ages 6 and 13 years). Analyses were adjusted for covariates in a step-wise manner. To assess whether the association between inflammatory exposures and child autistic traits differs depending on the child’s genetic liability, an interaction term for each exposure and child PGS ASD was added in separate linear mixed models. A significant interaction term indicated that the effect of the exposure on child autistic traits depended on the child’s genetic susceptibility for autism.

#### Sensitivity analyses

We conducted four sensitivity analyses. First, to understand the association with clinical autistic traits, the continuous child autistic trait scores were replaced by dichotomized scores into clinical versus subclinical categories in logistic mixed-effects models. Second, we investigated the timing of the inflammatory exposure by replacing the averaged cytokine index, CRP levels and fever count by the measurements obtained at each timepoint during pregnancy. Third, we assessed individual cytokine associations with child autistic trait scores. Fourth, we stratified for child sex to estimate sex-specific associations between exposures and child autistic traits.

Models were fit using the lme4 packages in R (30). P-values below alpha=0.05 were considered statistically significant. We applied a false discovery rate multiple testing correction to account for multiple testing within each exposure model (31). The analysis plan for this project was preregistered on December 11, 2024, and is available on OSF registries (osf.io/4afqe).

#### Covariates

Self-report questionnaires at enrollment were used to assess maternal covariates including age (continuous), education (primary, secondary or higher), national origin (Dutch, non-Dutch), parity (nulliparous, multiparous), tobacco use (no, smoked until pregnancy was known, continued smoking during pregnancy), alcohol use (no, drank until pregnancy was known, continued drinking during pregnancy occasionally, continued drinking during pregnancy frequently), substance use during pregnancy (yes/no), and pre-pregnancy BMI (continuous). In addition, analyses were adjusted for maternal psychopathology, which was measured using the global severity index (GSI) (continuous) from the self-report Brief Symptom Inventory (BSI) at enrollment, which represents an overall psychopathology score combining multiple symptom domains (32). A technical covariate includes gestational age of the exposure (weeks). Analyses with the MPS-CRP additionally included estimated cell type proportions based on a cord-blood reference panel (33) and sample plate to adjust for batch effects. Child covariates include child sex (male/female) obtained via medical registers and age at SRS assessment (continuous).

#### Missing data

Covariate data was missing for maternal factors pre-pregnancy BMI (14.6%), maternal psychopathology (13.3%), substance use (8.3%), tobacco use (8.2%), alcohol use (5.2%), education (2.8%), national origin (0.3%), and parity (0.2%). Missing covariate data was imputed using multiple imputation chained equations (40 imputed datasets, 100 iterations) using the ‘mice’ package in R.

## Results

A total of 5,075 mother-child pairs were included in the current study. Sample characteristics are shown in Table 1. A non-response analysis comparing the included and excluded sample is shown in Supplementary Table 1.

**Table 1.** Study population characteristics (N=5,075)

|  | Mean | SD | N | % |
| --- | --- | --- | --- | --- |
| *Maternal factors* |  |  |  |  |
| Maternal age (years) | 30.9 | 4.7 |  |  |
| Pre-pregnancy BMI (kg/m<sup>2</sup>) | 23.4 | 4.0 |  |  |
| Dutch maternal national origin |  |  | 3,059 | 60.5 |
| Maternal education level |  |  |  |  |
| Primary education |  |  | 326 | 6.6 |
| Secondary education |  |  | 2,020 | 40.9 |
| Higher education |  |  | 2,588 | 52.5 |
| Nulliparity |  |  | 2,071 | 40.9 |
| Psychopathology (GSI) | 0.3 | 0.3 |  |  |
| Smoking during pregnancy |  |  |  |  |
| Never smoked |  |  | 3,543 | 76.0 |
| Smoked until pregnancy was known |  |  | 416 | 8.9 |
| Continued smoking in pregnancy |  |  | 700 | 15.0 |
| Alcohol use during pregnancy |  |  |  |  |
| Never drank |  |  | 1,858 | 38.6 |
| Drank until pregnancy was known |  |  | 691 | 14.4 |
| Continued drinking occasionally |  |  | 1790 | 37.2 |
| Continued drinking frequently |  |  | 471 | 9.8 |
| Substance use during pregnancy |  |  | 295 | 6.3 |
| *Child factors* |  |  |  |  |
| Child age |  |  |  |  |
| First assessment | 6.2 | 0.5 |  |  |
| Second assessment | 13.6 | 0.4 |  |  |
| Child sex: female |  |  | 2,551 | 50.3 |
| SRS score |  |  |  |  |
| First assessment | 4.13 | 4.5 |  |  |
| Second assessment | 5.00 | 4.04 |  |  |

After adjusting for covariates, the cytokine index (β=-0.01, 95% CI = -0.03-0.02, p=0.703), CRP (β=0.00, 95% CI = -0.03-0.03, p=0.813), fever (β=0.02, 95% CI = -0.04-0.07, p=0.509), maternal polygenic score (PGS) for CRP (β=-0.01, 95% CI=-0.04-0.02, p=0.409) and the DNA methylation profile score (MPS) as a proxy of CRP (MPS-CRP) at birth (β=0.00, 95% CI=-0.04-0.04, p=0.991) were not significantly associated with child autistic traits (Figure 1). Assessment-specific analyses also showed no significant associations between inflammatory exposures and autistic traits assessed at child age 6 and 13 years (Figure 1). The significant unadjusted association between CRP and autistic traits was attenuated to non-significance after adjustment for maternal pre-pregnancy BMI (Supplementary Table 2). Maternal characteristics including increased psychopathology, tobacco use during pregnancy, higher pre-pregnancy BMI, younger maternal age at birth, lower education and non-Dutch national origin were associated with more autistic traits (Supplementary Figure 1). Obstetric complications (hypertensive disorders during pregnancy, preterm birth) and child characteristics (polygenic score for ASD, male sex) were also associated with increased autistic traits (Supplementary Figure 1).

**Table 2.** Interaction with child genetic risk of autism (PGS ASD) on the association between prenatal inflammatory exposures and child autistic traits.

| Inflammatory exposure | Interaction $\beta$ (95% CI) | 95% CI | p-value |
| --- | --- | --- | --- |
| Cytokine Index | -0.02 | -0.05; 0.01 | 0.175 |
| CRP | 0.02 | -0.01; 0.04 | 0.264 |
| Fever | -0.03 | -0.09; 0.04 | 0.406 |
| Polygenic score CRP | 0.01 | -0.02; 0.04 | 0.539 |
| Methylation profile score CRP | -0.05 | -0.11; 0.01 | 0.099 |

Note: all estimates adjusted for maternal covariates (age, education, national origin, parity, tobacco use, alcohol use, substance use, pre-pregnancy BMI, maternal psychopathology, gestational age at assessment) and child covariates (sex, age at assessment).

**Figure 1.**
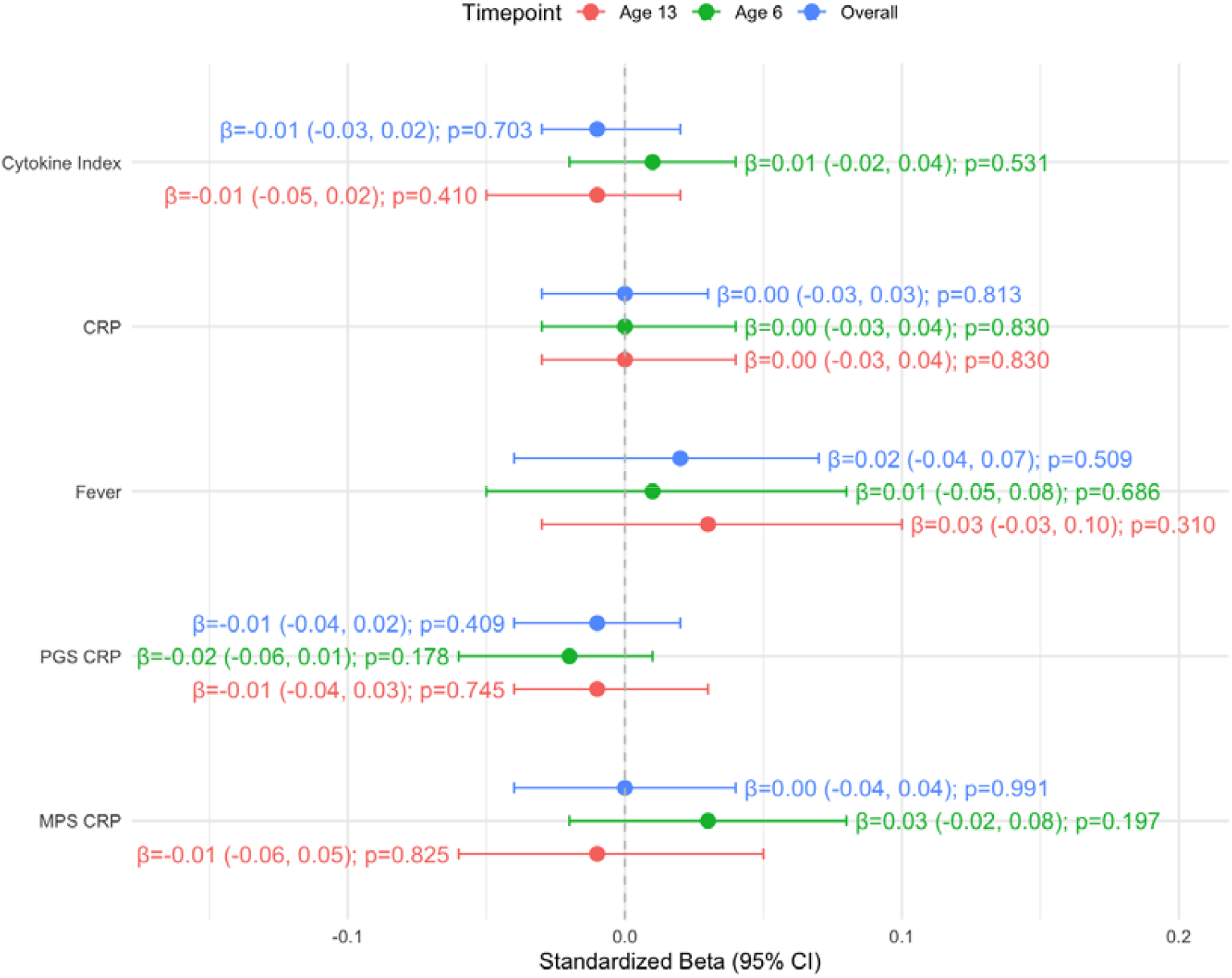
The association between prenatal inflammatory exposures and child autistic traits overall, and per assessment timepoint (child age 6 and 13 years). Adjusted for maternal covariates (age, education, national origin, parity, tobacco use, alcohol use, substance use, pre-pregnancy BMI, maternal psychopathology, gestational age at assessment) and child covariates (sex, age at assessment).

Next, we investigated if the associations between inflammatory exposures and child autistic traits were moderated by child genetic risk of autism using the polygenic score (PGS) for ASD. We found no evidence that the child PGS for ASD moderated the associations between the cytokine index, CRP, maternal fever during pregnancy, maternal PGS for CRP, and MPS-CRP at birth with child autistic traits (p>0.1) (Table 2).

We found no significant increased risk of clinical compared to subclinical autistic traits for any of the exposures (p>0.126) (Supplementary Table 3). Analyses per exposure timepoint (cytokine index and CRP at 13 and 20 weeks gestation; fever per trimester of pregnancy) also showed no association with child autistic traits (Supplementary Table 4). Individual cytokines were not significantly associated with child autistic traits (Supplementary Table 5). Stratification analyses showed no significant sex-specific effects, suggesting that the relationship between inflammatory exposures and child autistic traits is not different between boys and girls (Supplementary Table 6).

## Discussion

In this large, population-based longitudinal cohort, we found no evidence that detailed indicators of maternal immune activation – including maternal serum cytokine and CRP levels, self-reported maternal fever, maternal polygenic score for CRP and methylation profile score for CRP at birth – were associated with child autistic traits. Results were consistent across analyses stratified by timing of exposure, individual cytokines, child assessment age and for clinical versus subclinical autistic traits. Furthermore, incorporating the child’s polygenic score for ASD did not reveal any gene-environment interactions that conferred additional risk for autistic traits.

Our finding that maternal serum CRP during pregnancy was not associated with autistic traits is consistent with previous large population-based cohorts (n=788-4,165) (14–16). We also found no association with the maternal polygenic score for CRP, suggesting that neither serum CRP levels nor a genetic predisposition to chronically elevated CRP increases risk of autistic traits in the child. In contrast, a Finnish birth cohort (n=1,132) reported a 12% increased risk of childhood autism per unit increase in maternal CRP and a 40% increased risk for those in the top quintile (>5.84 mg/L) (34). Important differences with the current study include the use of clinical autism diagnosis, which may capture more severe phenotypes of neurodevelopmental disruption, and their lack of adjustment for maternal BMI, a well-established determinant of systemic CRP levels (17,35,36). In our analyses, the unadjusted association between maternal CRP and autistic traits was fully attenuated after adjustment for maternal BMI, suggesting that earlier studies may have captured the effect of maternal metabolic status rather than a direct inflammatory effect. Maternal BMI itself has been linked to ASD, with meta-analyses showing a dose-response relationship between BMI and autism risk in offspring (37–39), and we observed a similar positive association with autistic traits in our cohort. Large sibling-comparison studies suggest that familial factors contribute to this association (38). These findings highlight the importance of considering maternal BMI as both a confounder and a potential biological pathway in studies of maternal inflammation and child neurodevelopment.

We found no association between maternal serum cytokines and autistic traits. Evidence from smaller case-control studies is inconsistent and often undermined by limited sample sizes. One study of 84 ASD cases reported increased autism risk with higher mid-gestation levels of IFN-γ (OR = 1.52), IL-4 (OR = 1.51) and IL-5 (OR = 1.45), but no association for 14 other cytokines and minimal adjustment for confounding (12). A larger follow-up study of the same cohort (n=415) found no cytokines associated with ASD after adjustment compared to children with developmental delay and general population controls (13). They did find upregulated gestational GM-CSF, IL-1α, IL-6, and IFN-γ among children with ASD co-occurring with intellectual disability, suggesting etiological heterogeneity between ASD phenotypes (13). A nested case-control study (25 ASD cases) found an association with lower maternal IL-17A in early pregnancy, but not for seven other cytokines (IFN-γ, IL-16, Eotaxin, MCP-1, IL-1β, IL-8 and IL-6) (40). Taken together, robust evidence from large, well-adjusted population studies consistently points to the absence of a meaningful association between inflammatory biomarker levels and offspring autistic traits.

Similarly, we found no evidence that maternal fever during pregnancy was associated with child autistic traits, consistent with prior findings in this cohort (15). This contrasts with several large Swedish registry studies reporting increased autism risk (hazard ratio 1.16 (8); 30% increased odds (6); hazard ratio 1.79 (5)) following prenatal infection, and a meta-analysis of 36 epidemiological studies reporting a pooled 32% increase in odds of autism diagnosis (7). Interestingly, a Norwegian birth cohort reported increased risk of autism diagnosis only after fever in the second trimester, not in the first and third (41). Notably, these studies relied on clinically recorded infections, including inpatient diagnoses, which likely reflect more severe infections. This is supported by Danish registry data on 1.6 million children showing no increased risk of autism following maternal infection diagnosis, only in case of hospital admission for infection (hazard ratio 2.98) (42). These findings suggest that, unlike in preclinical models or high-risk case-control settings where inflammatory exposures are often severe and sustained, the modest inflammatory variation seen in the general population does not meaningfully impact autistic traits in childhood.

We additionally assessed the cord blood methylation profile score for CRP, which reflects stable epigenetic signatures of systemic inflammation (23,24). MPS approaches have been increasingly used in neurodevelopmental research, including ASD, where altered methylation signatures have been reported in post-mortem brain, placenta, and peripheral blood (43). We found no association between the MPS-CRP at birth and autistic traits. This finding aligns with earlier work in population-based cohorts reporting no direct link between MPS-CRP at birth (23,44), although indirect pathways were observed via cognitive function and sustained inflammation (44). These results suggest that mild inflammatory exposures detectable at the methylation level may not be sufficient in magnitude or duration to exert a pronounced impact on autistic traits later in life.

A central aim of our study was to evaluate whether child genetic liability amplified the risk of autistic traits following prenatal immune activation. We found no evidence that child genetic liability for ASD moderated the effects of inflammatory exposures on child autistic traits. This is consistent with findings from a sibling pair comparison in a large Swedish registry study, which found that the modest association between maternal infection and autism (HR 1.16) disappeared when comparing siblings discordant for exposure (HR 0.94), suggesting that shared genetic or familial factors may largely account for the observed association between maternal infection and autism (8). Overall, these results do not support a robust gene-environment interaction in which MIA disproportionately affects genetically susceptible children. In addition to common genetic risk captured by PGS, rare genetic variants may have large individual effects on autism risk, particularly in cases with co-occurring intellectual disability (45). Their potential role in moderating the effects of MIA warrant investigation in future work.

This study benefits from a large, population-based longitudinal design. A key strength is the consideration of and inclusion of multiple key confounders, exposures measured at different gestational timepoints and at birth (although the third trimester was not assessed), longitudinal, repeated measurements of autistic traits, and consideration of child genetic liability for ASD as a potential moderator. Limitations include the reliance on immune biomarkers measured in peripheral blood, which may not fully capture localized immune processes. Moreover, serum biomarkers likely reflect low-grade inflammation rather than acute infection-related spikes, given their short half-lives and timing of sample collection outside illness episodes. Fever was assessed via self-report, with no data on duration and severity. Finally, autistic traits were assessed using the social responsiveness scale, which does not capture categorical clinical diagnoses. Moreover, we did not differentiate between autism with and without co-occurring intellectual disability, which may represent distinct phenotypes. Despite these limitations, the cohort offers generalizable insights with sufficient power to detect meaningful prenatal risk factors for child autism (29) and ADHD symptoms (Zarchev et al., in press), as also reflected by the associations reported in this study between prenatal risk factors including maternal psychopathology, tobacco use during pregnancy, higher pre-pregnancy BMI, obstetric complications, higher polygenic score for ASD and male sex.

In summary, we found no evidence that maternal systemic cytokine or CRP levels, prenatal fever, genetic predisposition to elevated CRP, and a methylation proxy for CRP in cord blood were associated with autistic traits in childhood. These findings suggest that in the general population, typical fluctuations in maternal inflammation are unlikely to be a major driver of autistic traits. These findings may redirect the focus towards more severe inflammatory insults as well as other modifiable environmental influences and their interplay with genetic liability, which could offer potential targets for preventing adverse neurodevelopmental outcomes.

## Data Availability

All data produced in the present study are available upon reasonable request to the authors

## Notes

### Competing Interest Statement

The authors have declared no competing interest.

### Funding Statement

This study is supported by the NIH.

### Author Declarations

All participants provided written informed consent. The Medical Ethics Committee of Erasmus MC approved all study procedures.

