## Supplementary Files for "Investigating the association between maternal infection and inflammation and child autistic traits in a large population based cohort study"

### Supplementary Tables

**Supplementary Table 1.** Non-response analysis comparing included participants (N=5,075) to participants excluded from the analysis due to missing data on autistic traits (N = 2,723) or missing data on inflammation markers and fever (N = 2,047).

| | **Included in analysis (N=5,075)** | **Not in analysis (N=4,705)** | **Standardized mean difference** | **p-value** |
|---|---|---|---|---|
| *Maternal factors* | | | | |
| Maternal age (years), mean (SD) | 30.9 (4.7) | 28.8 (5.8) | 0.395 | <0.001 |
| Pre-pregnancy BMI (kg/m$^2$), mean (SD) | 23.4 (4.0) | 24.0 (4.8) | 0.135 | <0.001 |
| Dutch maternal national origin, N (%) | 3,059 (60.5%) | 1,486 (36.9) | 0.485 | <0.001 |
| Maternal education level, N (%) | | | 0.523 | <0.001 |
| Primary education | 326 (6.6) | 636 (17.6) | | |
| Secondary education | 2,020 (40.9) | 1,914 (52.8) | | |
| Higher education | 2,588 (52.5) | 1,073 (29.6) | | |
| Nulliparity, N (%) | 2,071 (40.9) | 2,518 (49.6) | 0.175 | <0.001 |
| Psychopathology (GSI), mean (SD) | 0.26 (0.33) | 0.39 (0.46) | 0.338 | <0.001 |
| Smoking during pregnancy, N (%) | | | 0.177 | <0.001 |
| Never smoked | 3,543 (76.0) | 2,512 (70.1) | | |
| Smoked until pregnancy was known | 416 (8.9) | 289 (8.1) | | |
| Continued smoking in pregnancy | 700 (15.0) | 783 (21.8) | | |
| Alcohol use during pregnancy, N (%) | | | 0.524 | <0.001 |
| Never drank | 1,858 (38.6) | 1,940 (62.6) | | |
| Drank until pregnancy was known | 691 (14.4) | 353 (11.4) | | |
| Continued drinking occasionally | 1790 (37.2) | 706 (22.8) | | |
| Continued drinking frequently | 471 (9.8) | 99 (3.2) | | |
| Substance use during pregnancy, N (%) | 295 (6.3) | 321 (9.1) | 0.102 | <0.001 |
| *Child factors* | | | | |
| Child age, mean (SD) | | | 0.024 | 0.503 |
| First assessment | 6.2 (0.5) | 6.2 (0.5) | | |
| Second assessment | 13.6 (0.4) | 13.6 (0.4) | | |
| Child sex: female, N (%) | 2,551 (50.3) | 2197 (48.3) | 0.040 | 0.055 |
| SRS score, mean (SD) | | | 0.006 | 0.887 |
| First assessment | 4.13 (4.5) | 4.47 (4.48) | | |
| Second assessment | 5.00 (4.04) | 5.02 (3.75) | | |
| *Inflammatory exposures* | | | | |
| CRP, mean (SD)[a] | 2.05 (1.35) | 2.26 (1.35) | 0.153 | <0.001 |
| IL-1β, mean (SD)[a] | 2.07 (0.70) | 2.01 (0.68) | 0.086 | 0.001 |
| IL-6, mean (SD)[a] | 0.96 (1.41) | 0.89 (1.42) | 0.050 | 0.063 |
| IL-17, mean (SD)[a] | 4.67 (0.80) | 4.60 (0.84) | 0.086 | 0.001 |
| IL-23, mean (SD)[a] | 10.25 (1.02) | 10.19 (1.02) | 0.062 | 0.021 |
| IFN-γ, mean (SD)[a] | 3.94 (0.82) | 3.86 (0.86) | 0.095 | <0.001 |
| Fever count, mean (SD) | 0.20 (0.51) | 0.14 (0.44) | 0.132 | <0.001 |

[a] Presented on log2 scale

**Supplementary Table 2.** The unadjusted association between prenatal inflammatory exposures and child autistic traits overall, and per assessment timepoint (child age 6 and 13 years).

| | **Overall** | | **Age 6** | | **Age 13** | |
|---|---|---|---|---|---|---|
| **Inflammatory exposure** | **β (95% CI)** | **P-value** | **β (95% CI)** | **P-value** | **β (95% CI)** | **P-value** |
| Cytokine Index | -0.018 | 0.166 | -0.011 | 0.458 | -0.019 | 0.235 |

| | | | | | | |
|---|---|---|---|---|---|---|
| | (-0.044; 0.008) | | (-0.042; 0.019) | | (-0.05; 0.012) | |
| CRP | **0.049 (0.023; 0.075)** | **<0.001** | **0.05 (0.022; 0.082)** | **0.001** | **0.044 (0.013; 0.075)** | **0.006** |
| Fever | 0.037 (-0.017; 0.091) | 0.209 | 0.034 (-0.03; 0.097) | 0.291 | 0.052 (-0.013; 0.117) | 0.117 |
| Polygenic score CRP | 0.017 (-0.010; 0.044) | 0.223 | 0.012 (-0.020; 0.044) | 0.457 | 0.017 (-0.016; 0.05) | 0.307 |
| Methylation profile score CRP | -0.038 (-0.075; 0.000) | 0.0523 | -0.034 (-0.076; 0.007) | 0.107 | -0.036 (-0.083; 0.011) | 0.130 |

**Supplementary Table 3.** The association between maternal immune activation prenatal inflammatory exposures and a dichotomized SRS score (clinical versus sub-clinical) in logistic mixed-effects model.

| **Inflammatory exposure** | **Odds ratio[1] (95% CI)** | **p-value** |
|---|---|---|
| Cytokine Index | -0.70 (-1.34; -0.06) | 0.310 |
| CRP | 0.22 (-0.32; 0.76) | 0.420 |
| Fever | -2.00 (-4.56; 0.56) | 0.126 |
| Polygenic score CRP | -0.322 (-0.915; 0.27) | 0.287 |
| Methylation profile score CRP | -0.113 (-0.834; 0.609) | 0.759 |

[1] Adjusted for maternal covariates (age, education, national origin, parity, tobacco use, alcohol use, substance use, pre-pregnancy BMI, maternal psychopathology, gestational age at assessment) and child covariates (sex, age at assessment).

**Supplementary Table 4.** The association between the cytokine index, CRP levels and fever per exposure timepoint with child autistic traits.

| **Inflammatory exposure** | **Timepoint** | **β (95% CI)[1]** | **P-value** |
|---|---|---|---|
| Cytokine index | 13 weeks gestation | -0.008 (-0.040; 0.024) | 0.623 |
| | 20 weeks gestation | -0.007 (-0.035; 0.022) | 0.655 |
| CRP | 13 weeks gestation | 0.007 (-0.027; 0.041) | 0.685 |
| | 20 weeks gestation | 0.002 (-0.029; 0.033) | 0.903 |
| Fever | Trimester 1 | 0.079 (-0.011; 0.17) | 0.088 |
| | Trimester 2 | -0.017 (-0.13; 0.097) | 0.767 |
| | Trimester 3 | -0.003 (-0.123; 0.119) | 0.973 |

[1] Adjusted for maternal covariates (age, education, national origin, parity, tobacco use, alcohol use, substance use, pre-pregnancy BMI, maternal psychopathology, gestational age at assessment) and child covariates (sex, age at assessment).

**Supplementary Table 5.** The association between individual cytokines and child autistic traits overall and per assessment timepoint (child age 6 and 13 years).

| **Cytokine** | **Overall** | | **Age 6** | | **Age 13** | |
|---|---|---|---|---|---|---|
| | **β (95% CI)[1]** | **P-value** | **β (95% CI)[1]** | **P-value** | **β (95% CI)[1]** | **P-value** |
| IL-1β | 0.0004 (-0.028; 0.028) | 0.976 | 0.013 (-0.021; 0.047) | 0.439 | -0.001 (-0.035; 0.032) | 0.947 |
| IL-6 | -0.007 (-0.035; 0.022) | 0.650 | 0.013 (-0.021; 0.046) | 0.460 | -0.017 (-0.051; 0.016) | 0.313 |
| IL17 | -0.027 (-0.056; 0.001) | 0.058 | -0.016 (-0.050; 0.018) | 0.359 | -0.030 (-0.064; 0.004) | 0.082 |

| | | | | | | |
|---|---|---|---|---|---|---|
| IL-23 | -0.0009 (-0.029; 0.027) | 0.951 | 0.001 (-0.032; 0.035) | 0.931 | -0.004 (-0.038; 0.029) | 0.780 |
| IFN-*y* | -0.018 (-0.046; 0.010) | 0.216 | 0.002 (-0.032; 0.035) | 0.927 | -0.030 (-0.064; 0.003) | 0.075 |

[1] Adjusted for maternal covariates (age, education, national origin, parity, tobacco use, alcohol use, substance use, pre-pregnancy BMI, maternal psychopathology, gestational age at assessment) and child covariates (sex, age at assessment).

**Supplementary Table 6.** Sensitivity analysis stratified for child sex to estimate sex-specific associations between prenatal inflammatory exposures and child autistic traits.

| **Inflammatory exposure** | **Child sex: Female (n=2,551)** | | **Child sex: Male (n=2,524)** | | **Interaction term (*childsex)** | |
|---|---|---|---|---|---|---|
| | **β (CI 95%)[1]** | **P-value** | **β (CI 95%)[1]** | **P-value** | **β (CI 95%)[1]** | **P-value** |
| Cytokine Index | -0.004 (-0.036; 0.028) | 0.804 | -0.011 (-0.058; 0.036) | 0.644 | 0.015 (-0.04; 0.071) | 0.604 |
| CRP | 0.000 (-0.002; 0.041) | 0.991 | 0.006 (-0.045; 0.057) | 0.813 | -0.012 (-0.068; 0.043) | 0.661 |
| Fever | -0.010 (-0.071; 0.052) | 0.760 | 0.049 (-0.041; 0.1139) | 0.287 | -0.068 (-0.175; 0.040) | 0.217 |
| Polygenic score CRP | -0.029 (-.063; 0.005) | 0.102 | 0.009 (-0.042; 0.060) | 0.724 | -0.041 (-0.101; 0.019) | 0.181 |
| Methylation profile score CRP | 0.002 (-0.041; 0.045) | 0.937 | -0.015 (-0.090; 0.059) | 0.688 | 0.011 (-0.073; 0.095) | 0.796 |

[1] Adjusted for maternal covariates (age, education, national origin, parity, tobacco use, alcohol use, substance use, pre-pregnancy BMI, maternal psychopathology, gestational age at assessment) and child covariates (sex, age at assessment).

**Supplementary Figure 1.** Associations between maternal and child characteristics and autistic traits. Standardized beta coefficients are shown. Asterisks indicate significance of the association.

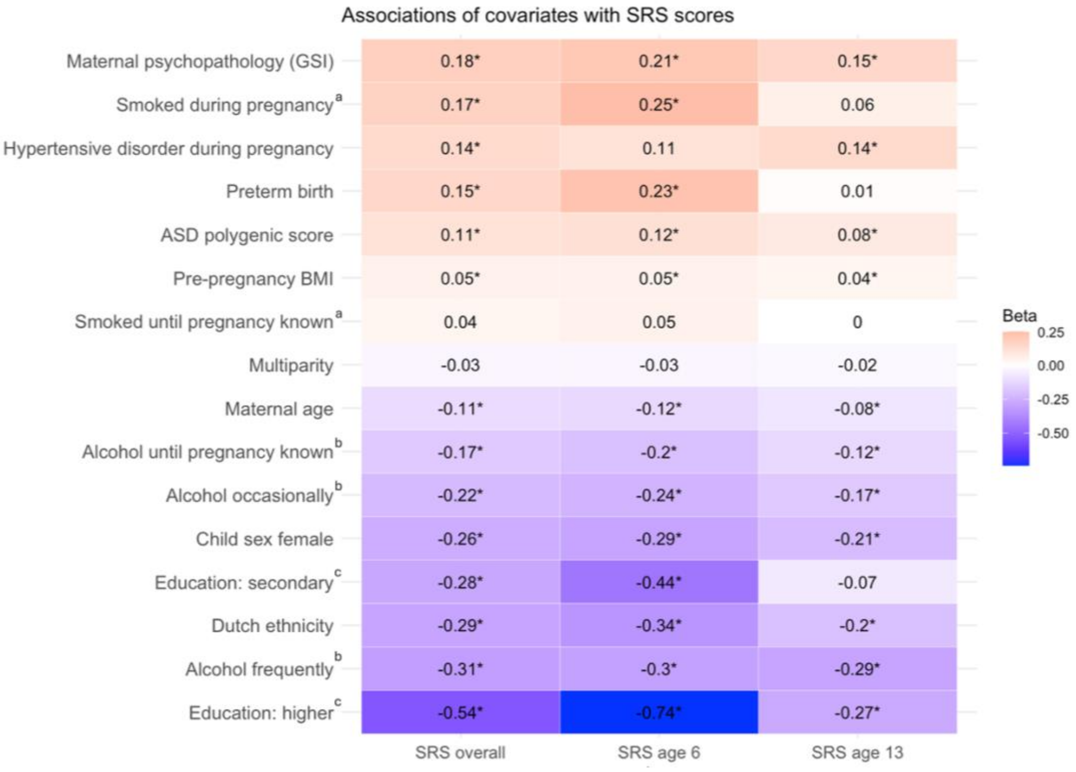


Legend: a=reference group no tobacco use, b= reference group no alcohol use; c= reference group primary education.
